# DeepDrug2: A Germline-focused Graph Neural Network Framework for Alzheimer’s Drug Repurposing Validated by Electronic Health Records

**DOI:** 10.1101/2025.05.13.25327570

**Authors:** Victor OK Li, Yang Han, Jacqueline CK Lam, Jocelyn Downey

**Author notes:** Equal contributions (Victor OK Li, Yang Han, Jacqueline CK Lam). Corresponding and senior authors: Victor OK Li and Jacqueline CK Lam. **Email:** and.

## Abstract

Alzheimer’s disease (AD) is a complex neurodegenerative disorder with limited therapeutic options. The original DeepDrug framework by Li et al. (2025) relied on somatic mutation data and emphasized long genes to guide AD drug repurposing. However, emerging evidence suggests that germline genetic variants play a more central role in AD pathogenesis. In response, we develop DeepDrug2, an enhanced AI-driven framework for AD drug repurposing centered on germline mutations and validated using real-world electronic health records (EHRs). DeepDrug2 introduces four major innovations. First, it proposes a different hypothesis prioritizing germline over somatic mutations in influencing AD risk. Second, it updates the signed directed heterogeneous biomedical graph by removing somatic mutations, long genes, and expert-led genes from the previous version, and incorporating new genes identified in recent genome-wide association study (GWAS) findings. Third, it generates a new list of drug candidates by encoding this updated graph into a new embedding space via a graph neural network (GNN) and calculating drug-gene scores. Fourth, it performs real-world clinical validation using EHR data from over 500,000 individuals (including more than 4,000 AD cases) in the UK Biobank, evaluating associations between drug usage and AD onset while controlling for demographic and comorbidity factors. DeepDrug2 has identified several promising drug candidates. Among the top 15 candidates with sufficient medication records to support statistically powered analysis, Amlodipine (a calcium channel blocker), Indapamide (a thiazide-like diuretic), and Atorvastatin (a statin) were significantly associated with reduced AD risk (*p* < 0.05). These findings highlight the role of germline mutations in guiding AD drug repurposing and emphasize the value of integrating real-world clinical data into AI-driven drug discovery. To further validate these candidates, future work will involve experimental studies using mouse and zebrafish models of AD. DeepDrug2 offers a promising strategy to support future clinical studies and expand therapeutic options for AD. Future work will evolve DeepDrug2 into a more powerful, versatile, and precise tool for AI-driven drug repurposing in neurodegenerative diseases by deeply integrating advanced LLM capabilities, prioritizing critical disease mechanisms, including tau pathology, and holistically incorporating multi-modal data sources.

## 1. Introduction

Alzheimer’s disease (AD) significantly deteriorates human health and quality of life, affecting approximately 50 million people globally. Despite decades of research, no effective disease-modifying treatment or preventive therapy for AD has been established. Drug development remains a lengthy and expensive process, requiring an estimated USD $2.6 billion to bring a single drug candidate to regulatory approval [1]. To date, only six drugs (Donepezil, Rivastigmine, Galantamine, Memantine, Aducanumab, and Lecanemab) and one drug combination comprising two of these six drugs (Donepezil and Memantine) have been approved by the US Food and Drug Administration (FDA) for AD treatment, with most therapies aimed at symptom management rather than modifying the underlying disease process [2].

Given these challenges, drug repurposing, i.e., identifying new therapeutic uses for existing approved drugs, has emerged as a promising strategy to accelerate drug discovery, offering a faster, lower-risk path to AD treatments [3, 4]. Computational methods have increasingly driven drug repurposing efforts through biomedical data. Early AD drug repurposing strategies primarily utilized statistical analyses of molecular data. A widely used technique is transcriptomic signature matching, which involves deriving differential gene expression signatures from AD versus control samples and identifying drugs whose perturbation profiles inversely correlate with the disease signature [5]. However, signature matching methods are limited by the quality and availability of gene expression data and often overlook disease mechanisms that operate beyond transcriptomic changes, particularly those embedded in molecular interaction networks. In contrast, network-based approaches emerged to model the complex molecular landscape of AD. These methods often integrate AD-associated genes, protein-protein interaction (PPI) networks, and drug-target data to assess the network proximity of drugs to disease modules [6]. However, traditional network proximity methods do not fully exploit the high-dimensional and non-linear relationships in biomedical networks.

Deep neural networks, particularly graph neural networks (GNNs), have recently been applied to model the complexity of biomedical systems for drug repurposing [7]. GNNs learn node representations (i.e., vector embeddings) by iteratively aggregating information from their neighbors, enabling the capture of both local and global structures within biomedical graphs. Biomedical entities, such as drugs, targets, pathways, and diseases, can be represented as nodes, with GNNs used to infer new edges, such as potential drug-disease associations, based on the learned representations. Several studies have tailored GNN models specifically for AD drug repurposing. NETTAG integrated genome-wide association study (GWAS) data and multi-omics evidence to predict AD risk genes and druggable targets through a GNN model designed to account for the sparse and modular topology of PPI networks [8]. A graph autoencoder model embedded nodes from an AD knowledge graph composed of drugs, genes, pathways, and gene ontology terms, facilitating the identification of AD drug candidates based on multi-level efficacy evidence, ranging from transcriptomic signatures to clinical trials [9]. DeepDrug presented the state-of-the-art advancements in AI-driven AD drug repurposing, incorporating expert-led domain-specific knowledge, such as long genes and somatic mutations detected in both blood and brain tissues [10–12], into a signed directed heterogeneous biomedical graph, enabling a GNN model to capture fine-grained drug-gene relationships and prioritize AD drug candidates validated through AD-related literature [13, 14].

However, despite its innovations in AI-driven AD drug repurposing, DeepDrug emphasized somatic mutations, which recent genetic and clinical research suggests are less central to sporadic (i.e., late onset) AD pathogenesis [15–18]. Large-scale GWAS studies have identified numerous germline variants significantly associated with AD risk, implicating core pathways involved in neuroinflammation, aging, and immune dysfunction [19–21]. These findings suggest that AD drug repurposing should prioritize germline-driven mechanisms, which better reflect the nature of AD pathophysiology, rather than relying predominantly on somatic mutation data. Furthermore, DeepDrug lacked validation using real-world clinical data, limiting its ability to assess whether the identified drug candidates can be translated into meaningful outcomes in patient populations.

In this study, we present DeepDrug2, an enhanced AI-driven framework for AD drug repurposing centered on germline mutations and validated using real-world electronic health records (EHRs). DeepDrug2 builds upon the strengths of the original DeepDrug but introduces four innovations (see Figure 1). First, it proposes a different hypothesis prioritizing germline over somatic mutations in influencing AD risk. Second, it updates the signed directed heterogeneous biomedical graph by removing somatic mutations, long genes, and expert-led genes from the previous version, and incorporating new genes identified in recent GWAS findings. Third, it generates a new list of drug candidates by encoding this updated graph into a new embedding space via a GNN model. Fourth, it performs real-world clinical validation using EHR data from over 500,000 individuals (including more than 4,000 AD cases) in the UK Biobank, evaluating associations between drug usage and AD onset while controlling for demographic and comorbidity factors.

**Figure 1.**
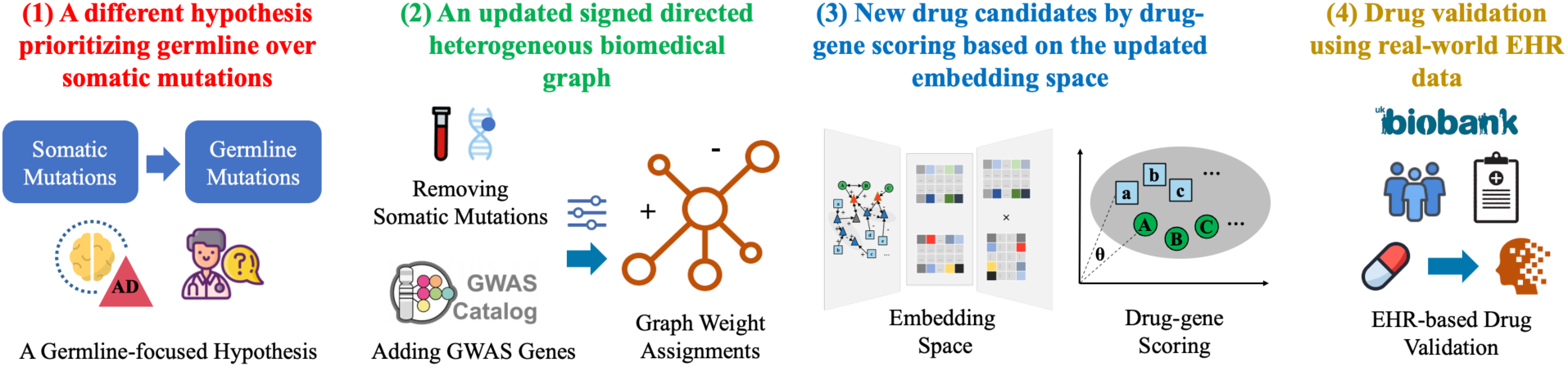
Novelties of DeepDrug2 for AD Drug Repurposing.

DeepDrug2 has identified several promising AD drug candidates. Among the top 15 candidates with sufficient medication records to support statistically powered analysis, Amlodipine (a calcium channel blocker), Indapamide (a thiazide-like diuretic), and Atorvastatin (a statin) were significantly associated with reduced AD risk (*p* < 0.05). These findings highlight the role of germline mutations in guiding AD drug repurposing and emphasize the value of integrating real-world clinical data into AI-driven drug discovery. To further validate these candidates, future work will involve experimental studies using mouse and zebrafish models of AD.

Looking ahead, future work will evolve DeepDrug2 into a more powerful, versatile, and precise tool for AI-driven drug repurposing in neurodegenerative diseases by deeply integrating the capabilities of advanced large language models (LLMs), prioritizing critical disease mechanisms, including tau pathology, and holistically incorporating multi-modal data sources. Fine-tuned LLMs trained on AD-relevant biomedical corpora will be used to generate new hypotheses and enhance the construction of the signed directed heterogeneous biomedical graph, enabling the assignment of node and edge weights based on semantic relevance and literature-derived evidence. Additionally, LLMs can support drug validation by predicting toxicity, pharmacokinetics, and clinical success probabilities through analysis of clinical trial reports, adverse event databases, and regulatory documentation. This integration of LLM-based inference with graph-based modeling positions DeepDrug2 to play a central role in the next generation of AI-driven frameworks for AD drug discovery.

## 2. Data and Methodology

Our proposed DeepDrug2 methodology has five steps. First, we collected relevant biomedical graph and clinical data (Section 2.1). Second, we constructed a heterogeneous directed biomedical graph encoding AD-related genes, proteins, drug targets, and drugs, capturing the core network properties of AD pathophysiology (Section 2.2). Third, we used a graph neural network to learn low-dimensional embeddings of drugs and genes, preserving the complex relationships inherent in the biomedical graph (Section 2.3). Fourth, we prioritized drug candidates based on the drug-gene scores calculated using the GNN-based embeddings (Section 2.4). Finally, we validated the selected drug candidates using EHRs to assess real-world clinical outcomes (Section 2.5). The details are described as follows.

### 2.1 Data Collection

We collected biomedical graph data on four node types, including genes, proteins, drug targets, and drugs, and three types of directed edges, including protein-protein interactions, drug-target interactions, and drug-drug interactions. Gene data were derived from a recent AD-focused GWAS study [20]. Drug and drug-drug interaction data were obtained from DrugBank [22]. Drug target information was aggregated from DrugBank [22], DrugCentral [23], ChEMBL [24], and BindingDB [25]. Protein-protein interaction and pathway data were sourced from the STRING database [26].

In addition to molecular and pharmacological data, we collected real-world clinical data from the UK Biobank to support post-hoc validation of drug candidates identified by DeepDrug2. The UK Biobank is a large-scale biomedical database containing health information from approximately 500,000 participants aged 40-69 years at recruitment [27]. For this study, we analyzed data from 502,163 individuals, including 4,451 participants diagnosed with AD. Medication usage data were extracted and mapped to the standardized drug names used in the biomedical graph, with brand name variations harmonized using DrugBank records [22]. AD diagnosis was based on ICD-9 and ICD-10 codes in hospital admission records. Clinical covariates, including age, gender, ethnicity, and key comorbidities (hypertension, diabetes, and coronary artery disease (CAD)) were extracted to control for potential confounding factors in statistical analysis [8].

### 2.2 Heterogeneous Signed Directed Biomedical Graph Construction

After data collection, we constructed a heterogeneous, signed, and directed biomedical graph to represent biologically meaningful interactions relevant to AD. The graph consisted of four distinct node types, including genes, proteins, drug targets, and drugs, and four types of directed edges, including gene-protein, protein-protein, drug-target, and drug-drug interactions. To improve computational efficiency and focus the analysis on biologically relevant structures, isolated nodes were removed from the final graph.

Following the methodology established in the DeepDrug framework [14], graph construction started from identifying key genes implicated in AD and approved drugs relevant to AD, guided by expert-led domain-specific knowledge. Drug targets bound to the selected drugs were then connected to the proteins corresponding to the identified genes through PPIs. The identified genes also underwent pathway enrichment analysis to determine their roles in AD-associated pathways in the PPI network. Finally, expert-guided knowledge was integrated into the biomedical graph by determining edge directions (e.g., whether one protein acts on another protein) and signs (e.g., whether a drug activates or inhibits its target) and assigning weights to nodes and edges (e.g., gene nodes weighted by their importance to AD based on statistical significance from GWAS analysis).

The original biomedical graph developed in DeepDrug [14] was further updated to reflect the updated hypothesis of AD pathophysiology, emphasizing germline over somatic mutations in influencing AD risk. The three major changes are listed as follows:

1. Removal of somatic mutation data: All somatic mutation features, long genes, and previously expert-curated gene sets focusing on somatic alterations were excluded based on recent GWAS evidence highlighting the predominance of germline mutations in contributing to AD risk [20].
2. Integration of germline-based AD genes: We incorporated 107 AD-associated genes identified from the recent GWAS findings [20]. Each gene node was weighted by its normalized –log(*p*) value to reflect the strength of GWAS association.
3. Updated pathway enrichment analysis: Using the STRING pathway analysis tool [26], we identified 11 statistically significant AD-related pathways (FDR < 0.05) from the KEGG database [28] based on the 107 AD-associated genes. For PPI edges involving proteins in these pathways, edge weights were further increased based on normalized –log(FDR) values to emphasize biologically relevant pathways in the graph.

The resulting graph captures the hierarchical flow from AD-associated genes through proteins and drug targets to candidate drugs, enabling advanced graph representation learning for further analysis. Figure 2 displays a simplified visualization of the updated DeepDrug2 biomedical graph, highlighting interactions among the top 50 nodes from each node category and showing the largest connected component in the graph.

**Figure 2.**
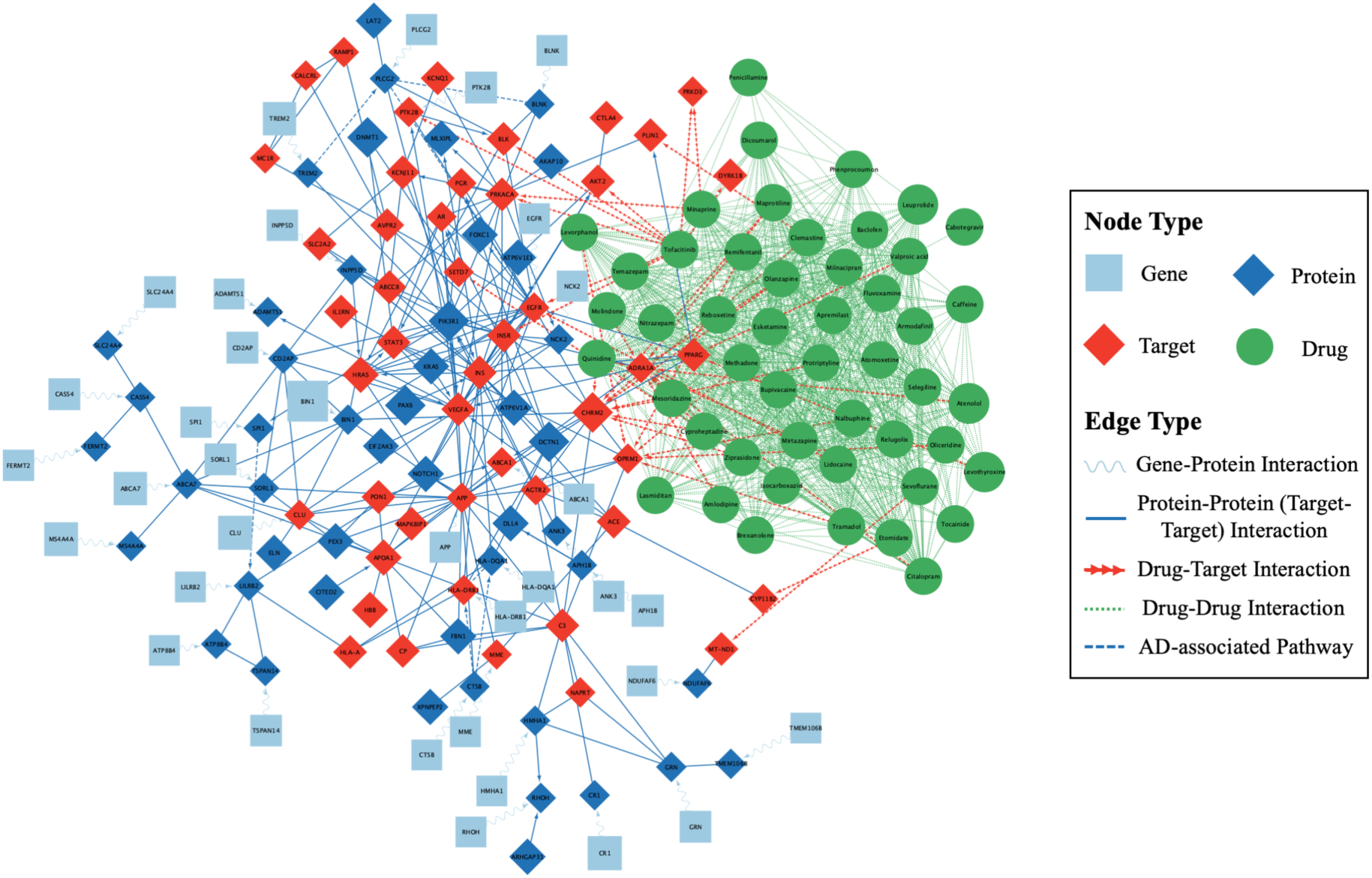
DeepDrug2 Heterogeneous Signed Directed Biomedical Graph (Simplified Version)

### 2.3 Graph Neural Network Model Development

After the biomedical graph construction, we used the signed directed GNN model proposed in the DeepDrug framework [14] to embed biomedical nodes, including genes and drugs, into a unified low-dimensional embedding space. The GNN framework is based on a graph autoencoder (GAE) architecture, which learns node embeddings by encoding both node features and the signed directed adjacency structure of the biomedical graph. Specifically, the encoder uses signed and directed graph convolutional network (GCN) layers to generate positive and negative embeddings for each node, which are then combined into source and target embeddings. The decoder reconstructs the network by optimizing an edge probability matrix derived from these embeddings. Conceptually, the GNN framework is described as follows:

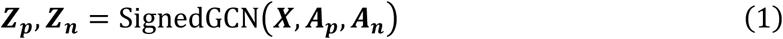

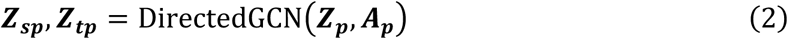

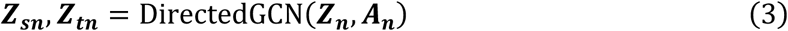

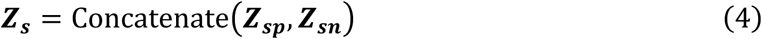

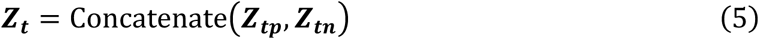

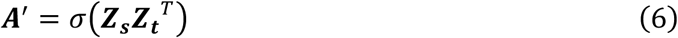

where

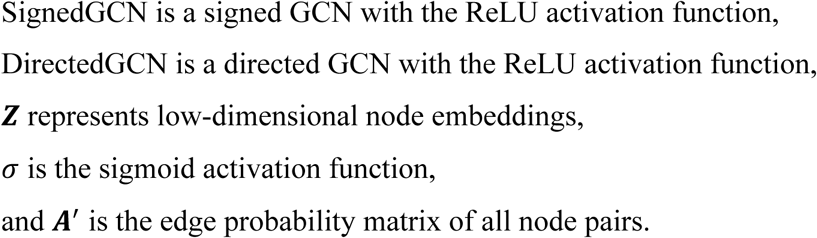

The GNN model was trained using a link prediction task, framing the problem as binary classification to estimate the likelihood of an edge between any two nodes, and enabling drug candidates to be prioritized based on their proximity to AD-associated genes. To enhance biological relevance, pathway-guided regularization was applied. This included two auxiliary classification losses based on node embeddings: (1) node type classification (cross-entropy loss), ensuring each node maps to one of the four predefined types (gene, protein, drug target, drug) and (2) pathway membership classification (binary cross-entropy loss), allowing nodes to belong to zero or more pathway, preserving the network structure and the functional convergence/divergence of AD-related pathways in the embedding space.

### 2.4 Drug Candidate Scoring and Selection

Following model training and validation, we used the trained GNN to generate embedding vectors for all drug and gene nodes in the biomedical graph. To prioritize drug candidates, we computed drug-gene scores based on the proximity of each drug to AD-associated genes in the embedding space. A higher score indicated a drug was more closely related to AD genes. Incorporating the direction and sign of biomedical interactions, we quantified drug-gene relationships based on the anti-correlation of embeddings, prioritizing drug inhibitors close to AD-risk genes and drug activators close to AD-protective genes:

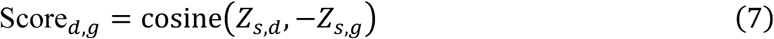

where

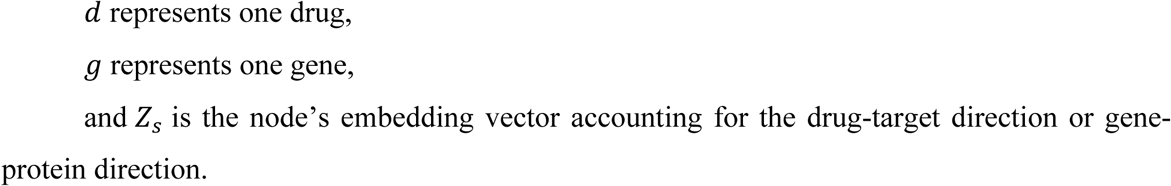

Next, we calculated the score of a single drug candidate based on the average drug-gene scores across all AD-related genes:

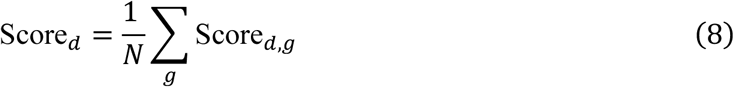

where

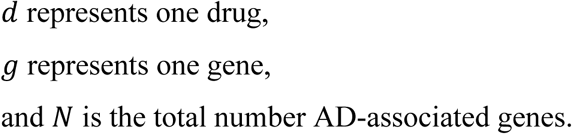

Further, we selected the top *K* single drug candidates based on their drug-gene scores. These candidates were subsequently validated using real-world EHR data.

### 2.5 Drug Validation Using Electronic Health Records

To assess the real-world effectiveness of the drug candidates identified by DeepDrug2, we performed post-hoc validation using EHR data from the UK Biobank. Specifically, we employed a logistic regression model to evaluate the association between candidate drug usage and the risk of AD. In this model, AD diagnosis served as the binary outcome variable, while the primary exposure variable was binary drug usage (1 = exposed; 0 = unexposed), indicating whether an individual had a documented medication history of using the drug. To account for potential confounding effects, the model included six demographic and clinical covariates, including age, gender, ethnicity, and comorbidities known to be associated with AD risk, including hypertension, diabetes, and CAD [8]. Each candidate drug was evaluated independently. To ensure statistical robustness, we included only those drugs for which at least 70 individuals were classified as exposed, following the rule of thumb that regression analysis requires a minimum of ten observations per independent variable. Conceptually, the statistical model is described as follows:

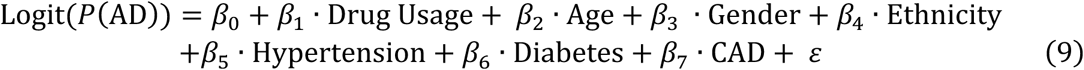

where

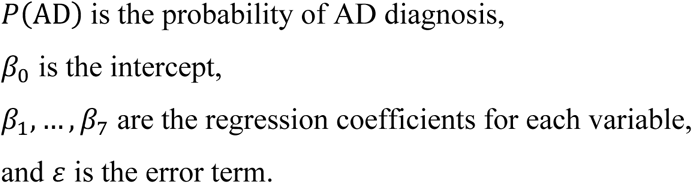

The coefficient β_1,_ represents the estimated effect of drug exposure on AD risk, controlling for age, gender, ethnicity, and comorbidities. Specifically, β_1,_ quantifies the log odds change in the probability of an AD diagnosis associated with exposure to a specific drug, while holding all other covariates constant. To facilitate interpretation, β₁ was exponentiated to obtain the odds ratio (OR): OR = *e*^β1^. If OR > 1, drug exposure increases the risk of AD. If OR < 1, drug exposure decreases the risk of AD. If OR = 1, drug exposure has no effect on AD risk.

## 3. Results

### 3.1 Experimental Setup

We used a random 80/10/10 split of all existing edges on the graph as the training set, validation set, and test set. The training set was used for the GNN model training. The validation set was used for hyperparameter selection and early stopping during training. After model training, the test set was used for performance evaluation. The optimized node embeddings were used to classify the existence of edges on the test set, using the Area under the ROC Curve (AUC) and Average Precision (AP) as the model performance evaluation metrics, which are two commonly used evaluation metrics for link prediction [29]. The AUC and AP metrics measure how well the model can distinguish existing edges and non-existent edges on the biomedical graph. AUC ranges from 0 to 1, with an AUC of 1 indicating perfect performance. AP is the area under the precision-recall curve, summarizing the trade-off between precision and recall at various thresholds into a single value. AP ranges from 0 to 1, with an AP of 1 indicating perfect performance.

The hyperparameter settings are listed as follows. The dimension of node feature vectors was 260, including four dimensions from the expert-guided node weight vector (representing gene, protein, drug, and target nodes) and 256 dimensions derived from the corresponding biological structure (gene sequence, protein sequence, or drug compound). Each GCN in the encoder had two layers, and the hidden unit size was 128. The embedding dimension size was selected empirically and set to 32. The number of training epochs was 100. The Adam optimizer was used to learn the model parameters, with a learning rate of 0.01. For the directed GCNs, two hyperparameters α and β were used ([0.4, 0.6], [0.5, 0.5], or [0.6, 0.4]) to control the relative importance of source and target information. The weight of the pathway-guided regularization term was 0.1, 0.5, or 1.0.

### 3.2 Performance Evaluation

As shown in Figure 3, the signed directed GNN model demonstrates exceptional performance, achieving 98% classification accuracy, with an AUC score of 0.98 and an AP score of 0.98. These high metrics underscore the model’s ability to effectively capture and represent the complex biomedical graph structure, accurately modeling the relationships between drug and gene nodes. This performance indicates that the model is not only robust in its ability to classify drug-gene interactions but also highly proficient in preserving the intricate dependencies and interactions inherent to AD-related molecular networks.

**Figure 3.**
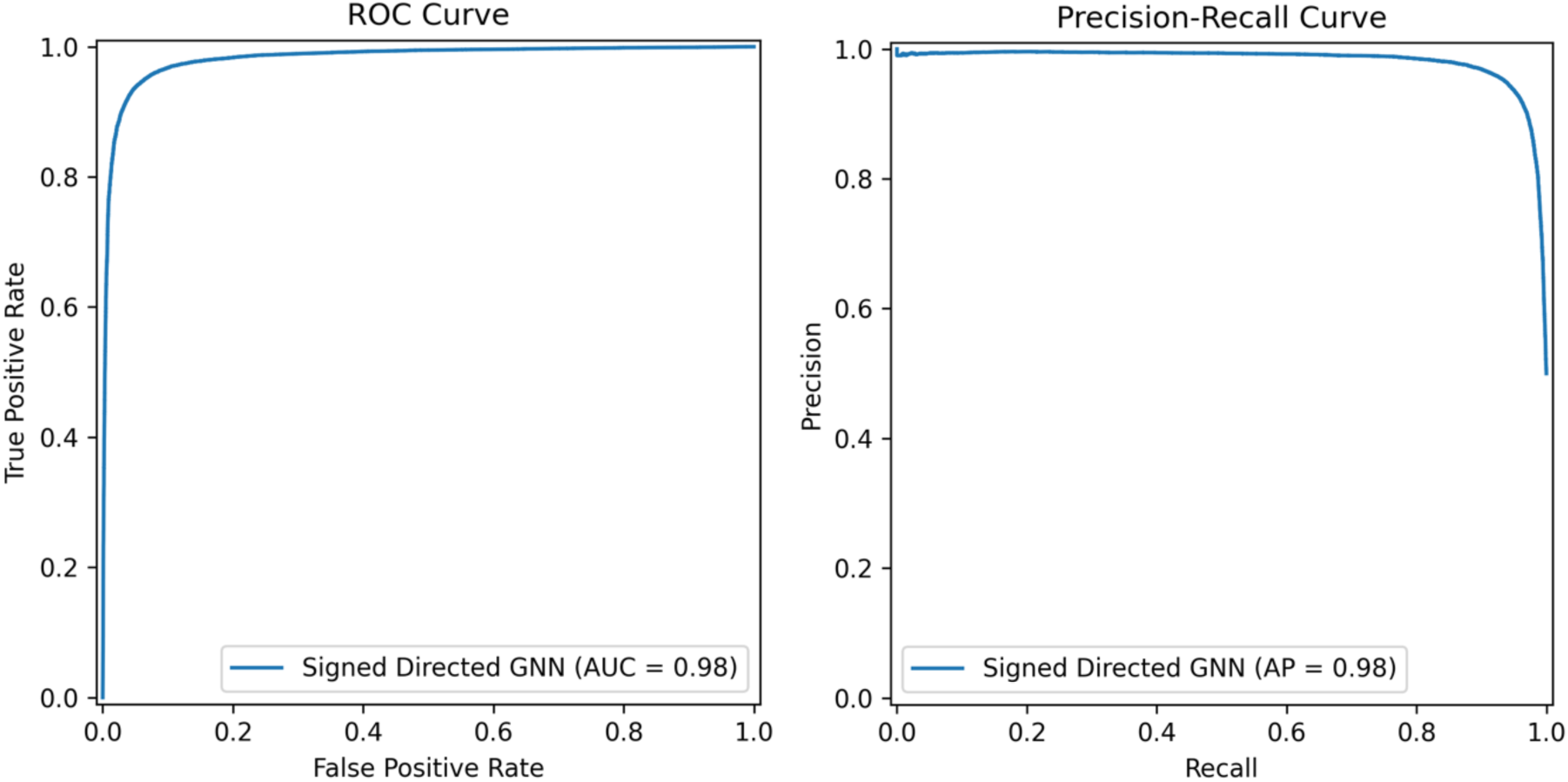
ROC and Precision-recall Curves of the GNN Model.

### 3.3 Top Drug Candidates

We generated a list of top repurposed approved drugs based on the DeepDrug2 framework. Table 1 shows the top 50 drug candidates. Compared to the top 15 drug candidates identified by the original DeepDrug framework [14], DeepDrug2 retains 10 of the original 15 top candidates, indicating significant but not complete overlap. These shared candidates include Tofacitinib, Raloxifene, Empagliflozin, Doxercalciferol, Palbociclib, Febuxostat, Olanzapine, Baricitinib, Miconazole, and Valproic Acid. Notably, Tofacitinib, Baricitinib, Empagliflozin, and Doxercalciferol were also part of the lead five-drug combination identified by the original DeepDrug framework [14].

**Table 1.**
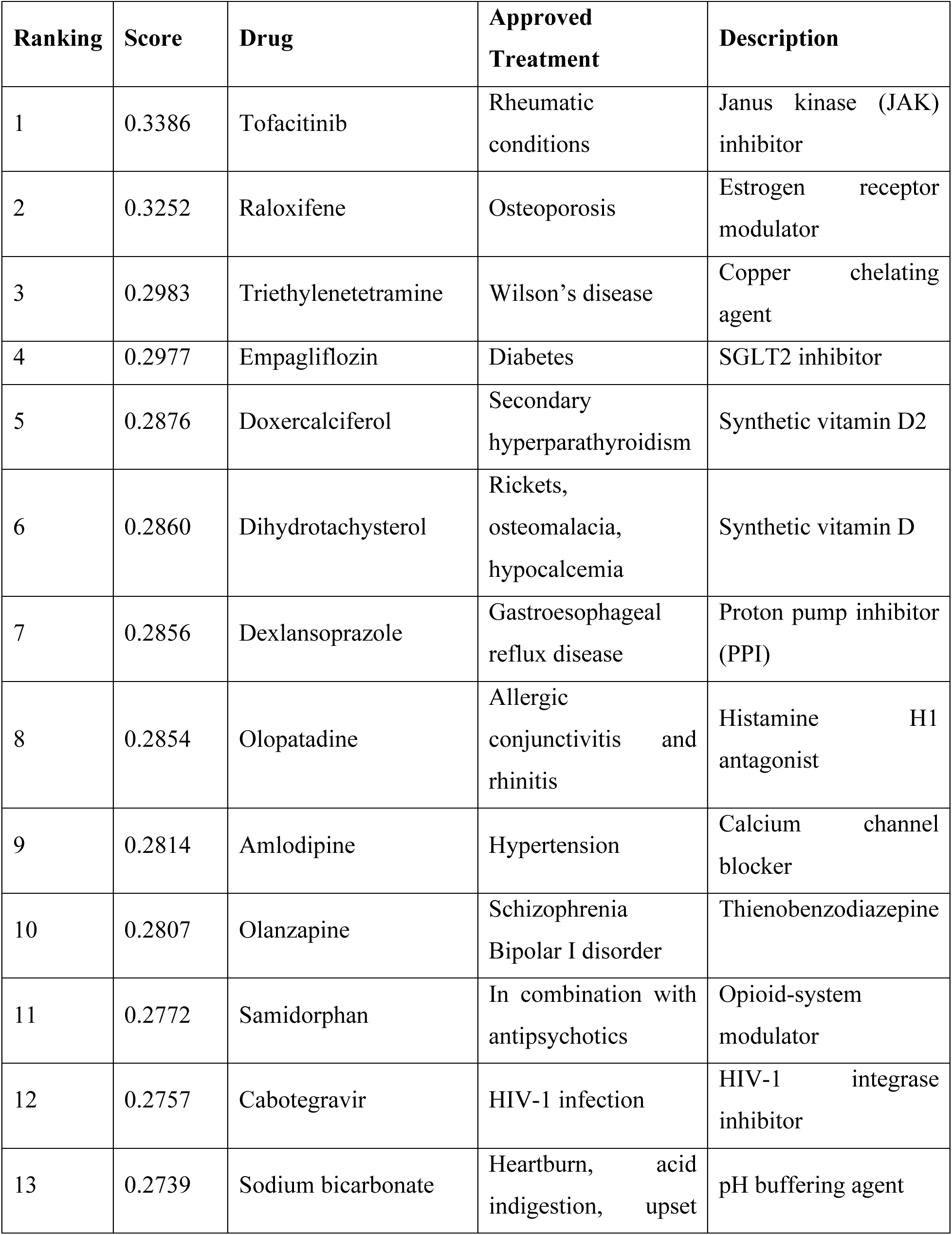
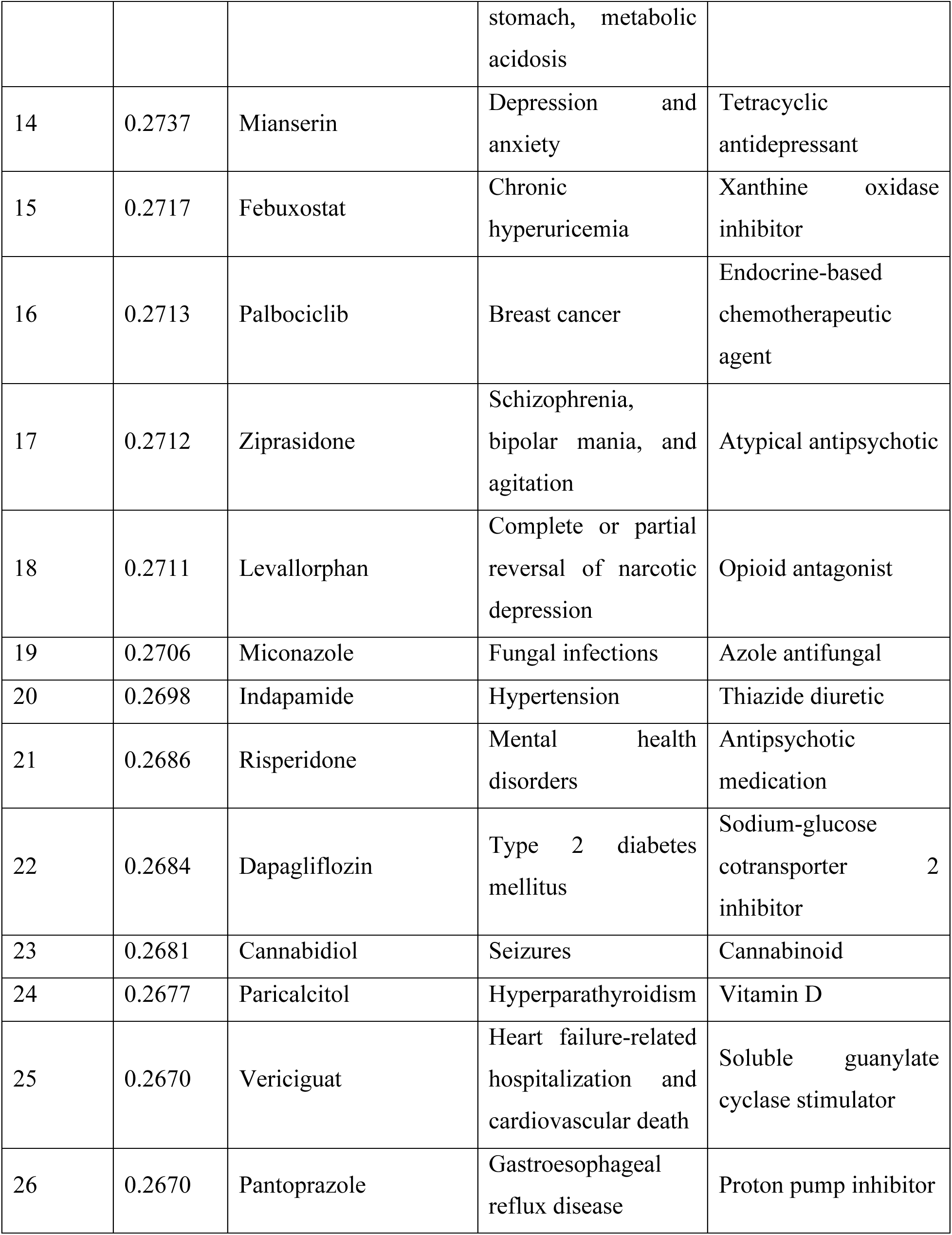
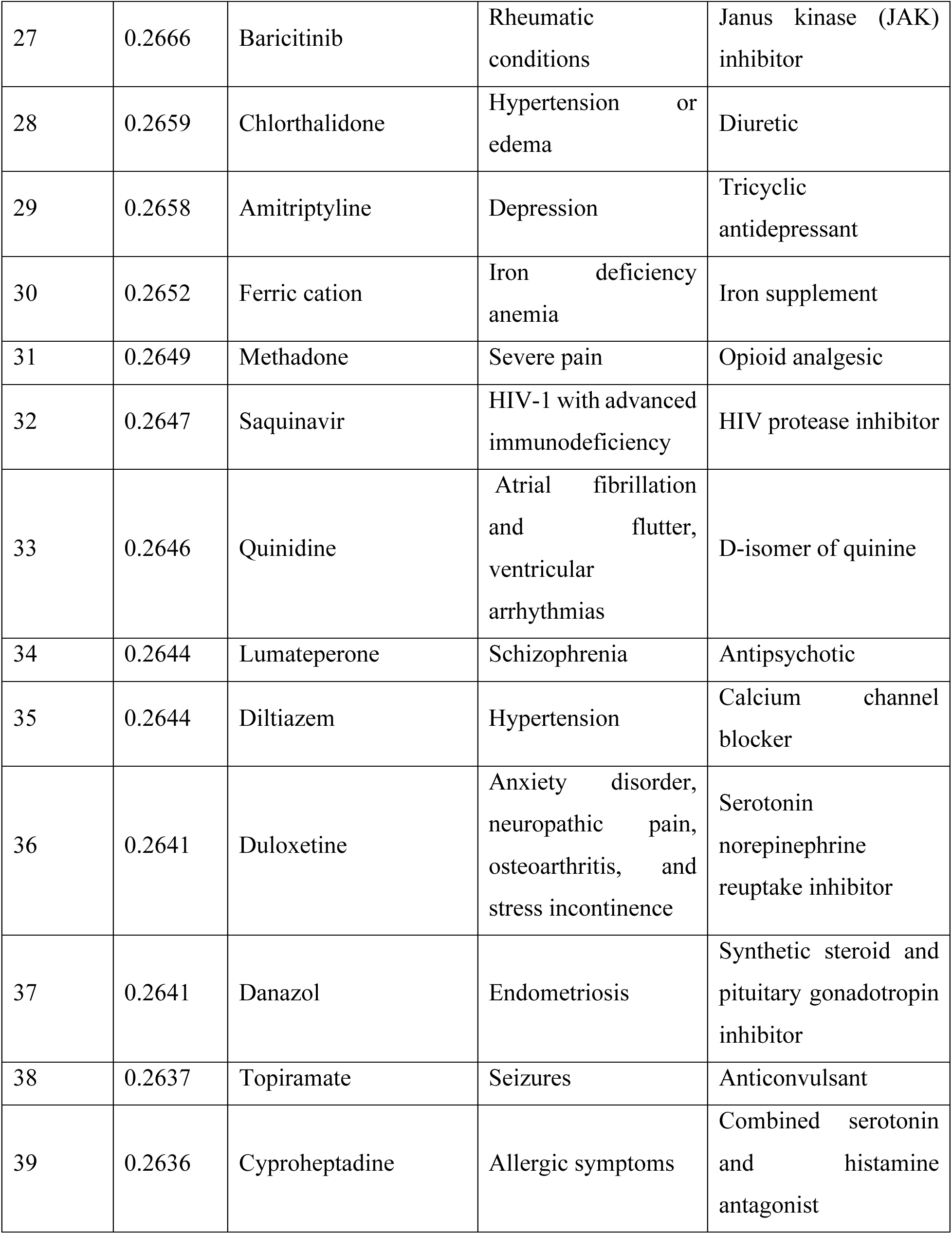
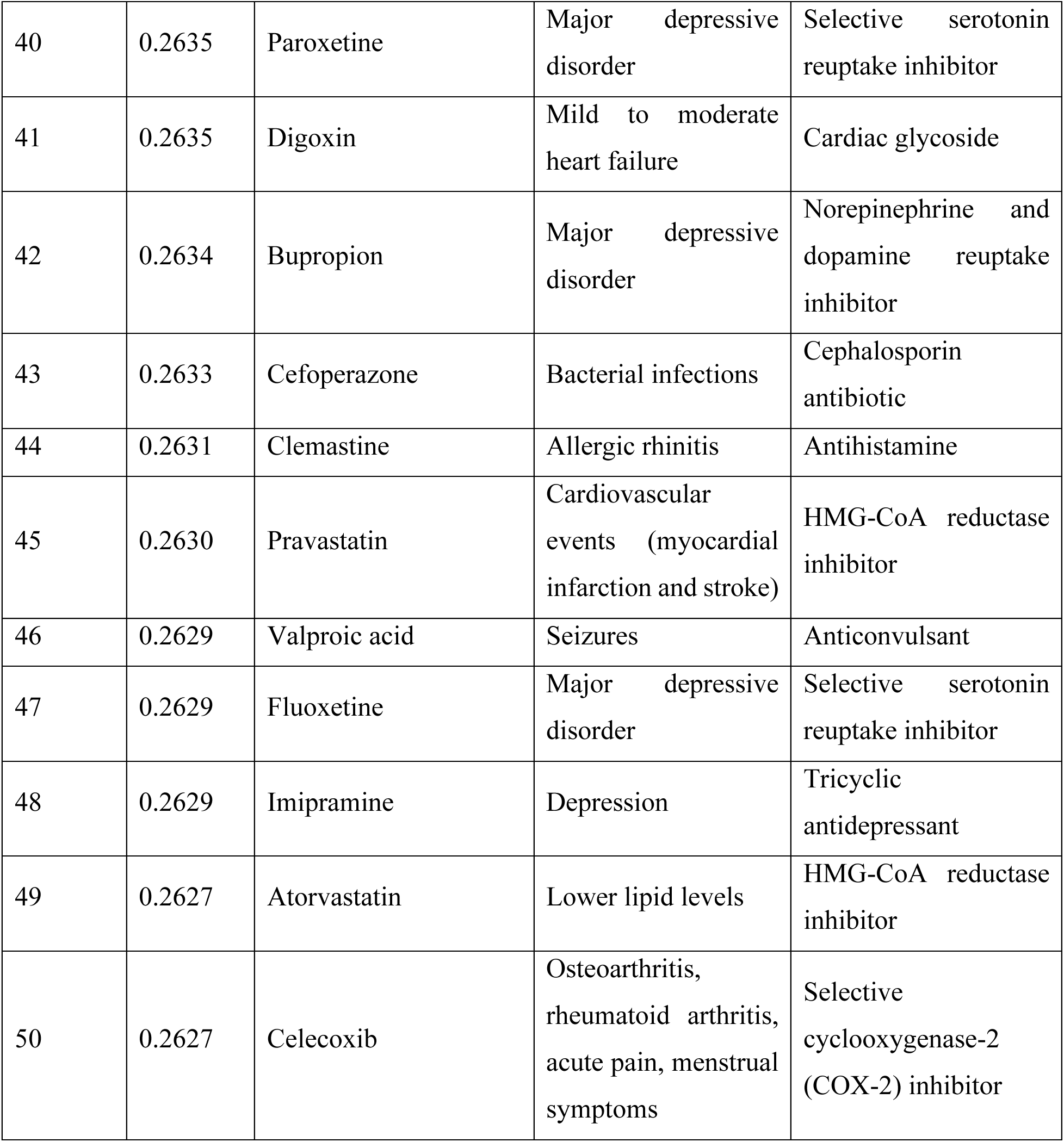
Top Repurposed Drug Candidates for AD Identified by DeepDrug2.

| Ranking | Score | Drug | Approved Treatment | Description |
| --- | --- | --- | --- | --- |
| 1 | 0.3386 | Tofacitinib | Rheumatic conditions | Janus kinase (JAK) inhibitor |
| 2 | 0.3252 | Raloxifene | Osteoporosis | Estrogen receptor modulator |
| 3 | 0.2983 | Triethylenetetramine | Wilson's disease | Copper chelating agent |
| 4 | 0.2977 | Empagliflozin | Diabetes | SGLT2 inhibitor |
| 5 | 0.2876 | Doxercalciferol | Secondary hyperparathyroidism | Synthetic vitamin D2 |
| 6 | 0.2860 | Dihydrotachysterol | Rickets, osteomalacia, hypocalcemia | Synthetic vitamin D |
| 7 | 0.2856 | Dexlansoprazole | Gastroesophageal reflux disease | Proton pump inhibitor (PPI) |
| 8 | 0.2854 | Olopatadine | Allergic conjunctivitis and rhinitis | Histamine H1 antagonist |
| 9 | 0.2814 | Amlodipine | Hypertension | Calcium channel blocker |
| 10 | 0.2807 | Olanzapine | Schizophrenia<br>Bipolar I disorder | Thienobenzodiazepine |
| 11 | 0.2772 | Samidorphan | In combination with antipsychotics | Opioid-system modulator |
| 12 | 0.2757 | Cabotegravir | HIV-1 infection | HIV-1 integrase inhibitor |
| 13 | 0.2739 | Sodium bicarbonate | Heartburn, acid indigestion, upset stomach, metabolic acidosis | pH buffering agent |
| 14 | 0.2737 | Mianserin | Depression and anxiety | Tetracyclic antidepressant |
| 15 | 0.2717 | Febuxostat | Chronic hyperuricemia | Xanthine oxidase inhibitor |
| 16 | 0.2713 | Palbociclib | Breast cancer | Endocrine-based chemotherapeutic agent |
| 17 | 0.2712 | Ziprasidone | Schizophrenia, bipolar mania, and agitation | Atypical antipsychotic |
| 18 | 0.2711 | Levallorphan | Complete or partial reversal of narcotic depression | Opioid antagonist |
| 19 | 0.2706 | Miconazole | Fungal infections | Azole antifungal |
| 20 | 0.2698 | Indapamide | Hypertension | Thiazide diuretic |
| 21 | 0.2686 | Risperidone | Mental health disorders | Antipsychotic medication |
| 22 | 0.2684 | Dapagliflozin | Type 2 diabetes mellitus | Sodium-glucose cotransporter 2 inhibitor |
| 23 | 0.2681 | Cannabidiol | Seizures | Cannabinoid |
| 24 | 0.2677 | Paricalcitol | Hyperparathyroidism | Vitamin D |
| 25 | 0.2670 | Vericiguat | Heart failure-related hospitalization and cardiovascular death | Soluble guanylate cyclase stimulator |
| 26 | 0.2670 | Pantoprazole | Gastroesophageal reflux disease | Proton pump inhibitor |
| 27 | 0.2666 | Baricitinib | Rheumatic conditions | Janus kinase (JAK) inhibitor |
| 28 | 0.2659 | Chlorthalidone | Hypertension or edema | Diuretic |
| 29 | 0.2658 | Amitriptyline | Depression | Tricyclic antidepressant |
| 30 | 0.2652 | Ferric cation | Iron deficiency anemia | Iron supplement |
| 31 | 0.2649 | Methadone | Severe pain | Opioid analgesic |
| 32 | 0.2647 | Saquinavir | HIV-1 with advanced immunodeficiency | HIV protease inhibitor |
| 33 | 0.2646 | Quinidine | Atrial fibrillation and flutter, ventricular arrhythmias | D-isomer of quinine |
| 34 | 0.2644 | Lumateperone | Schizophrenia | Antipsychotic |
| 35 | 0.2644 | Diltiazem | Hypertension | Calcium channel blocker |
| 36 | 0.2641 | Duloxetine | Anxiety disorder, neuropathic pain, osteoarthritis, and stress incontinence | Serotonin norepinephrine reuptake inhibitor |
| 37 | 0.2641 | Danazol | Endometriosis | Synthetic steroid and pituitary gonadotropin inhibitor |
| 38 | 0.2637 | Topiramate | Seizures | Anticonvulsant |
| 39 | 0.2636 | Cyproheptadine | Allergic symptoms | Combined serotonin and histamine antagonist |
| 40 | 0.2635 | Paroxetine | Major depressive disorder | Selective serotonin reuptake inhibitor |
| 41 | 0.2635 | Digoxin | Mild to moderate heart failure | Cardiac glycoside |
| 42 | 0.2634 | Bupropion | Major depressive disorder | Norepinephrine and dopamine reuptake inhibitor |
| 43 | 0.2633 | Cefoperazone | Bacterial infections | Cephalosporin antibiotic |
| 44 | 0.2631 | Clemastine | Allergic rhinitis | Antihistamine |
| 45 | 0.2630 | Pravastatin | Cardiovascular events (myocardial infarction and stroke) | HMG-CoA reductase inhibitor |
| 46 | 0.2629 | Valproic acid | Seizures | Anticonvulsant |
| 47 | 0.2629 | Fluoxetine | Major depressive disorder | Selective serotonin reuptake inhibitor |
| 48 | 0.2629 | Imipramine | Depression | Tricyclic antidepressant |
| 49 | 0.2627 | Atorvastatin | Lower lipid levels | HMG-CoA reductase inhibitor |
| 50 | 0.2627 | Celecoxib | Osteoarthritis, rheumatoid arthritis, acute pain, menstrual symptoms | Selective cyclooxygenase-2 (COX-2) inhibitor |

DeepDrug2 preserves core candidates identified by DeepDrug, showing continuity in prioritizing JAK inhibitors (e.g., Tofacitinib and Baricitinib) and metabolic regulators (e.g., Empagliflozin and Doxercalciferol). However, it substantially broadens the candidate space by introducing drugs from more diverse therapeutic classes (e.g., antidepressants and statins). The absence of several DeepDrug candidates reflects DeepDrug2’s updated biomedical graph, which removes somatic mutation data and incorporates germline mutations and pathways, leading to a more comprehensive drug prioritization strategy.

### 3.4 Drug Validation Results

Among the top 50 drug candidates identified by DeepDrug2 (Table 1), 15 had sufficient medication records in the UK Biobank to enable statistical validation using EHR data. We assessed the association between exposure to each drug and the risk of AD using logistic regression, adjusting for age, gender, ethnicity, and key comorbidities. The results are summarized in Table 2.

**Table 2.** Top Repurposed Drug Candidates Validated by EHR Data.

| Ranking<sup>1</sup> | Drug<sup>2</sup> | Odds Ratio (OR) | *p*-value<sup>3</sup> |
| --- | --- | --- | --- |
| 2 | Raloxifene | 0.71 | 0.4962 |
| 9 | Amlodipine | 0.87 | 0.0182 * |
| 10 | Olanzapine | 1.62 | 0.2491 |
| 20 | Indapamide | 0.41 | 0.0006 *** |
| 21 | Risperidone | 2.16 | 0.1328 |
| 26 | Pantoprazole | 1.49 | 0.0787 |
| 35 | Diltiazem | 1.05 | 0.8351 |
| 36 | Duloxetine | 0.69 | 0.5163 |
| 38 | Topiramate | 1.13 | 0.8640 |
| 40 | Paroxetine | 2.12 | 0.0000 *** |
| 45 | Pravastatin | 0.91 | 0.5258 |
| 47 | Fluoxetine | 1.63 | 0.0001 *** |
| 48 | Imipramine | 1.05 | 0.9583 |
| 49 | Atorvastatin | 0.81 | 0.0007 *** |
| 50 | Celecoxib | 0.77 | 0.5604 |
| Notes: |  |  |  |
| 1. Ranking among the top 50 drug candidates identified by DeepDrug2. |  |  |  |
| 2. Only 15 drugs with sufficient medication records in the UK Biobank were selected. |  |  |  |
| 3. *: $p < 0.05$; **: $p < 0.01$; ***: $p < 0.001$ | | | |

Several candidate drugs demonstrated statistically significant associations with AD risk. Notably, Indapamide (OR = 0.41, *p* = 0.0006) and Atorvastatin (OR = 0.81, *p* = 0.0007) were significantly associated with a reduced risk of AD, supporting their potential as AD drugs. Amlodipine, a calcium channel blocker, also showed a modest but statistically significant protective effect (OR = 0.87, *p* = 0.0182). These findings align with previous studies suggesting the neuroprotective effects of certain antihypertensive and lipid-lowering agents in reducing AD risk [30–32].

Conversely, several drugs were associated with increased AD risk. Most notably, Paroxetine (OR = 2.12, *p* < 0.0001) and Fluoxetine (OR = 1.63, *p* = 0.0001) exhibited strong associations with higher AD risk. Both are selective serotonin reuptake inhibitors (SSRIs). While SSRIs have been explored for their potential neurocognitive effects [33], these results suggest the need for further investigation into long-term SSRI exposure and its impact on cognitive decline.

Other drugs, including Olanzapine, Risperidone, and Pantoprazole, showed elevated odds ratios but did not reach statistical significance, potentially due to limited sample size or confounding by indication. It is also worth noting that some drugs, such as Raloxifene, Duloxetine, and Celecoxib, trended toward protective associations but lacked statistical power to confirm significance.

In addition to the 15 drugs analyzed, many of the top-ranking candidates identified by DeepDrug2 could not be evaluated using the UK Biobank data due to insufficient exposure records. This limitation reflects the relatively low prescription rates or recent approval status of some drugs in the general population. As a result, the current validation may underrepresent the full therapeutic potential of the DeepDrug2 candidates. Future validation efforts leveraging larger or more diverse real-world datasets will be critical to systematically evaluate these additional candidates.

## 4. Discussion

DeepDrug2 introduces significant methodological advancements compared to previous AI-driven approaches for AD drug discovery, leveraging a signed directed GNN to model complex molecular interactions within a biomedical knowledge graph. Compared to the original DeepDrug framework, DeepDrug2 shifts the focus explicitly toward germline mutations, integrating emerging evidence underscoring the central role of hereditary genetic variations in AD susceptibility. Specifically, it constructs a specialized biomedical graph devoid of somatic mutations and long genes, incorporating recent GWAS-identified germline variants. Moreover, this graph is transformed via a GNN into a refined embedding space, uniquely prioritizing germline genetic factors in drug-gene scoring. The model achieved strong classification performance (AUC = 0.98, AP = 0.98), underscoring its ability to capture nuanced relational patterns between drug and gene nodes in the AD molecular network. These results affirm the model’s capacity to represent both the directionality and polarity of interactions, providing a more biologically grounded and informative platform for prioritizing drug candidates.

Compared to the original DeepDrug framework, DeepDrug2 preserves much of the foundational insight while improving and expanding the candidate space. Ten of the top 15 candidates from DeepDrug were retained in DeepDrug2’s top 50 list, demonstrating continuity in highlighting JAK inhibitors (e.g., Tofacitinib and Baricitinib) and metabolic regulators (e.g., Empagliflozin and Doxercalciferol). This overlap validates the consistency of the core methodology while also highlighting how updates to the underlying graph, such as the removal of somatic mutations and the incorporation of germline genetics, can meaningfully shift the prioritization landscape. Importantly, DeepDrug2 introduces greater diversity in therapeutic classes, surfacing antidepressants, antiepileptics, statins, and calcium channel blockers, thereby broadening the horizon of potential AD therapeutics.

Further innovation is evident in DeepDrug2’s real-world clinical validation using extensive EHRs from over half a million individuals in the UK Biobank, rigorously evaluating drug usage against AD onset. This clinical validation strategy surpasses theoretical predictions common in previous methodologies, yielding statistically robust associations between drugs and reduced AD risk. Of the 50 top-ranked candidates, 15 had sufficient medication records to permit statistical evaluation. Among these, three drugs, Indapamide, Atorvastatin, and Amlodipine, were significantly associated with reduced AD risk after adjusting for confounding variables. These results echo prior epidemiological studies suggesting protective effects of antihypertensive and lipid-lowering therapies on cognitive health, reinforcing the potential repurposing value of these widely used agents. Conversely, some candidate drugs were associated with an increased risk of AD. Paroxetine and Fluoxetine, both SSRIs, showed significant positive associations with AD diagnosis. While SSRIs have been studied for their neuropsychiatric benefits in aging populations, these findings call attention to potential long-term risks and underscore the need for more nuanced investigations into their role in neurodegeneration. It is also important to note that many top-ranked DeepDrug2 candidates were excluded from the EHR validation due to limited real-world exposure data. For example, drugs like Tofacitinib and Empagliflozin, despite high prioritization scores, are relatively new or prescribed for narrow indications, resulting in insufficient representation in the UK Biobank cohort. This highlights an inherent limitation of retrospective validation in existing datasets and signals the need for complementary validation strategies using more diverse, contemporary, or longitudinally rich health records.

DeepDrug2 advances the AI-driven drug discovery landscape by effectively bridging genetic insights with clinical outcomes, streamlining drug repurposing pipelines while improving prediction accuracy. Unlike many existing approaches that focus primarily on identifying drug targets from large molecular libraries, DeepDrug2 adopts a patient-centric strategy, prioritizing drugs through the integration of genetic data and EHRs. This alignment with AD pathology ensures predictions are not only mechanistically grounded but also clinically relevant. By incorporating EHR-based validation using patient-specific data, DeepDrug2 enhances both its real-world applicability and predictive precision. This strategic synthesis of genetic and clinical data marks DeepDrug2 as a distinctive and forward-looking model in the AI-driven search for effective AD therapies.

Overall, DeepDrug2 presents a significant advancement in AI-driven drug repurposing for AD. Its focus on germline mutations and expanded candidate space provides a strong foundation for future translational studies. The findings from clinical validation of DeepDrug2’s predictions underscore the importance of integrating real-world data into computational drug repurposing pipelines. Future work will benefit from broader EHR data integration and prospective validation to better understand the causal effects of these candidate drugs on AD progression. Translating these insights into clinical interventions will be a critical step toward realizing the therapeutic potential of DeepDrug2 in AD prevention and treatment.

## 5. Future Development

The following strategic directions outline the envisioned innovations that will distinguish DeepDrug2 from existing AI-driven drug repurposing approaches, solidifying its position as a cutting-edge platform for therapeutic discovery in neurodegenerative diseases.

### 5.1 LLM-based Drug Repurposing

LLMs have shown great promise in accelerating drug repurposing by generating prioritized candidate lists informed by extensive biomedical literature. In the context of AD, recent studies have validated ChatGPT-generated AD drug candidates using real-world EHR data [34], and have further improved LLM-guided discovery by integrating structured AD-related knowledge (e.g., from Gene Ontology and DrugBank) into prompt engineering strategies for LLaMA-based models [35]. These approaches underscore the utility of general-purpose LLMs in rapidly synthesizing biomedical evidence to identify and prioritize plausible therapeutic candidates for AD.

Beyond general-purpose models, recent advancements in domain-specific LLMs have demonstrated competitive performance in predicting drug-related properties. For example, DeepMind’s Tx-LLM and TxGemma [36, 37], fine-tuned on hundreds of diverse therapeutic tasks across chemical and biological modalities, have been utilized to predict disease-associated genes, drug-target binding affinity, toxicity, and clinical trial success, enabling integrated, end-to-end drug discovery workflows. Further, specialized fine-tuning has led to models such as AD-GPT, designed specifically for AD-related information retrieval tasks, including the identification of relationships among genes, brain regions, and disease pathology [38]. However, current LLM applications remain largely disconnected from graph-based drug repurposing frameworks and often lack fine-tuning for AD-specific data tailored for drug discovery.

Future work can focus on developing fine-tuned LLMs trained on AD-relevant chemical and biomedical corpora to improve understanding of molecular structures, disease mechanisms, and bioactivity profiles. Such models could systematically extract structured biomedical knowledge from unstructured text, enabling the generation of new hypotheses and the discovery of latent links among genes, pathways, drug interactions, and disease progression. For the DeepDrug2 framework, this presents a compelling opportunity to enrich the biomedical graph with semantically meaningful nodes and edges derived from LLM-based inference. LLMs, with their ability to encode and synthesize massive amounts of biomedical literature, can be used to assign edge weights (e.g., drug-target interactions and protein-protein interactions) by evaluating co-occurrence patterns, semantic similarity, and literature-derived evidence scores. Similarly, node-level weights (e.g., genes and drugs), can be refined using LLM-inferred relevance to AD-specific pathology. The integration of LLMs into the construction of the signed directed heterogeneous biomedical graph can enable a more nuanced and biologically grounded representation of biomedical entities. This fusion of graph-based modeling with natural language-derived insights can support more comprehensive candidate prioritization in AD drug repurposing.

Moreover, LLMs hold promise for advancing drug validation, particularly in predicting toxicity profiles, clinical trial success probability, and pharmacokinetic characteristics, which are critical bottlenecks in drug discovery. By fine-tuning LLMs on datasets such as clinical trial reports, FDA drug labels, and adverse event databases, these models can provide text-based predictive signals regarding the safety and clinical viability of repurposed candidates. Incorporating these LLM-derived validation metrics into the DeepDrug2 framework can facilitate a more holistic and clinically informed candidate assessment process.

### 5.2 Targeting Tau Removal

Given the repeated clinical failures of amyloid-targeting therapies, there is increasing recognition that tau pathology may be a more proximal driver of AD [39]. This shift in therapeutic focus has fuelled interest in tau as a critical drug target [40]. Several AI-driven AD studies have begun to explore tau-related mechanisms. For instance, a knowledge graph-based machine learning framework was developed to predict gene candidates associated with tau pathology, offering a promising strategy for identifying novel drug targets [41]. Additionally, LLM-based multi-agent drug design pipelines have included specific use cases focused on generating candidate molecules that inhibit GSK-3β, a kinase known to promote tau hyperphosphorylation [42]. These developments highlight the potential of AI-driven approaches to accelerate tau-targeted drug discovery and underscore the opportunity to incorporate tau-specific biomarkers and pathways into AD therapeutics. However, current AI-driven drug repurposing frameworks have yet to fully integrate tau-related molecular and clinical data, limiting their capacity to precisely model tau-driven disease mechanisms and predict effective interventions.

Future work can focus on extending DeepDrug2 to incorporate tau-specific molecular features and pathway-level annotations, enabling more targeted identification of candidate compounds that modulate tau pathology. This could include integrating imaging and proteomic data, such as tau biomarker profiles, into the biomedical graph to better capture the mechanistic underpinnings of tau aggregation and spread. Furthermore, LLMs fine-tuned on tau-relevant biomedical literature could be leveraged to generate novel hypotheses, extract latent mechanistic patterns, and propose candidate interventions. Coupling these advances with experimental validation or EHR-based outcome modelling would help prioritize therapies most likely to yield clinical benefit, particularly in patient subgroups defined by tau burden or progression stage (e.g., Braak staging). As AI-driven drug repurposing platforms increasingly align molecular mechanisms with clinical endpoints, such tau-focused extensions of DeepDrug2 could play a pivotal role in addressing one of the most pressing gaps in AD therapeutic development.

### 5.3 Multi-modal Data Integration

AD involves complex interactions across genomics, proteomics, imaging, and clinical domains. Advanced AI methods have begun to harness multi-modal data for modelling these molecular and phenotypic interactions. For example, ATOMICA utilizes a universal graph-based modelling approach, integrating diverse data types, such as proteomics, metabolomics, and transcriptomics, to enhance predictive power and uncover novel biological insights [43]. Additionally, structural biology tools, such as AlphaFold [44], have provided accurate three-dimensional protein models that, when combined with multi-omics data, can refine predictions of drug-target interactions [45, 46]. Furthermore, recent efforts have constructed heterogeneous AD knowledge graphs by using LLMs to extract relationships between features in different biomedical datasets, such as imaging and gene expression data [47]. Despite these advances, the potential of LLMs to systematically synthesize and integrate information across diverse biomedical modalities remains underutilized in multi-modal AD drug repurposing frameworks.

Future work can focus on leveraging LLMs fine-tuned on multi-modal biomedical corpora, such as omics annotations, neuroimaging reports, and clinical narratives, to generate rich, semantically informed embeddings that complement structured datasets. By integrating LLM-derived insights with graph-based representations (e.g., biomedical knowledge graphs enriched with multi-omics and imaging data), DeepDrug2 can achieve a more holistic understanding of AD pathology. This LLM-augmented multi-modal integration has the potential to uncover novel genotype-phenotype relationships and improve the biological interpretability of drug predictions, ultimately accelerating the identification and prioritization of therapies tailored to the heterogeneity of AD.

## 6. Conclusion

DeepDrug2 advances AD drug repurposing by integrating a signed directed GNN with an updated biomedical graph that shifts the foundational hypothesis: from emphasizing somatic mutations to focusing on germline genetic risks and pathways. This conceptual shift reflects a more current understanding of AD pathophysiology and enables a biologically grounded, generalizable approach to AD drug candidate prioritization. DeepDrug2 demonstrates strong predictive performance and uncovers a broader, more diverse array of therapeutic candidates. Real-world validation supports the clinical relevance of several top drug candidates, reinforcing their translational potential. Future work will evolve DeepDrug2 into a more powerful, versatile, and precise tool for AI-driven drug repurposing in neurodegenerative diseases by deeply integrating advanced LLM capabilities, prioritizing critical disease mechanisms, including tau pathology, and holistically incorporating multi-modal data sources. Future work will also involve experimental studies in mouse and zebrafish models of AD to provide critical preclinical evidence for advancing the identified candidates toward therapeutic development.

## Acknowledgments

ChatGPT-4o was used to improve the language of the manuscript and to assist in the writing of the manuscript (only covering literature review and discussion of results). The authors reviewed the content generated and took full responsibility for the content of the manuscript.

We would like to thank Prof. David Rubinsztein for the helpful discussions and valuable insights regarding targeting tau removal in AD drug development. We would also like to thank Shanshan Wang for her research assistance and contribution to the discussion and future development sections. This research was supported in part by the US National Academy of Medicine (NAM) Healthy Longevity Catalyst Award 2021 and “An Artificial Intelligence (AI)-driven Causal Approach for Early Diagnosis and Treatment of Late Onset Alzheimer’s Disease” under Seed Fund for Collaborative Research, The University of Hong Kong, June 2023.

## Data Availability

The drug and drug-drug interaction data were obtained from DrugBank (https://go.drugbank.com). The drug target data were obtained from DrugBank (https://go.drugbank.com), DrugCentral (https://drugcentral.org), ChEMBL (https://www.ebi.ac.uk/chembl/), and BindingDB (https://www.bindingdb.org/). The protein-protein interaction and pathway data were obtained from STRING (https://string-db.org). The preprocessed biomedical network datasets, including genes and their weights (Table S1), drugs and their weights (Table S2), proteins/targets and their weights (Table S3), and edges and their weights (Tables S4-7), are available in Supplementary Data. The UK Biobank data used in this study can be accessed by qualified researchers through formal application at https://www.ukbiobank.ac.uk, subject to approval and compliance with ethical guidelines.

